# Characterization of CSF inflammatory markers after hemorrhagic stroke and their relationship to disease severity

**DOI:** 10.1101/2023.12.05.23299566

**Authors:** Jessica Magid-Bernstein, Jennifer Yan, Alison L. Herman, Zili He, Conor W. Johnson, Hannah Beatty, Rachel Choi, Sofia Velazquez, Eitan Neeman, Guido Falcone, Jennifer Kim, Nils Petersen, Emily J. Gilmore, Charles Matouk, Kevin Sheth, Lauren Sansing

**Author notes:** Correspondence to: Jessica Magid-Bernstein, MD, PhD, Division of Neurocritical Care and Emergency Neurology, Department of Neurology, Yale School of Medicine, 100 York Street, Suite 1N, New Haven, CT 06511.

## Abstract

**Background:** The inflammatory response within the central nervous system is a key driver of secondary brain injury after hemorrhagic stroke, both in patients with intracerebral hemorrhage (ICH) and aneurysmal subarachnoid hemorrhage (aSAH). In this study, we aimed to characterize inflammatory molecules in the blood and cerebrospinal fluid (CSF) of patients within 72 hours of hemorrhage to understand how such molecules vary across disease types and disease severity.

**Methods:** Biological samples were collected from patients admitted to a single-center Neurosciences Intensive Care Unit with a diagnosis of ICH or aSAH between 2014 and 2022. Control CSF samples were collected from patients undergoing CSF diversion for normal pressure hydrocephalus. A panel of immune molecules in the plasma and CSF samples was analyzed using Cytometric Bead Array assays. Clinical variables, including demographics, disease severity, and intensive care unit length of stay were collected.

**Results:** Plasma and/or CSF samples were collected from 260 patients (188 ICH patients, 54 aSAH patients, 18 controls). C-C motif chemokine ligand-2 (CCL2), interleukin-6 (IL-6), granulocyte-colony stimulating factor (G-CSF), interleukin-8 (IL-8), and vascular endothelial growth factor (VEGF), were detectable in the CSF within the first 3 days after hemorrhage, and all were elevated compared to plasma. Compared with controls, CCL2, IL-6, IL-8, G-CSF, and VEGF were elevated in the CSF of both ICH and aSAH patients (p<0.01 for all comparisons). VEGF was increased in ICH patients compared to aSAH patients (p<0.01). CCL2, G-CSF, and VEGF in the CSF were associated with more severe disease in aSAH patients only.

**Conclusions:** Within 3 days of hemorrhagic stroke, proinflammatory molecules can be detected in the CSF at higher concentrations than in the plasma. Early concentrations of some pro-inflammatory molecules may be associated with markers of disease severity.

## INTRODUCTION

Despite significant advances in the acute management of hemorrhagic stroke and its devastating consequences, short– and long-term outcomes remain poor. Mortality remains as high as 40% after intracerebral hemorrhage (ICH) and 35% after aneurysmal subarachnoid hemorrhage (aSAH), and less than 40% of patients regain functional independence.^1–5^ The inflammatory response driven by the presence of blood within the brain parenchyma, ventricles, and subarachnoid space is a key driver of secondary brain injury.^11–15^

In animal models of ICH, multiple phases of inflammation occur within the brain.^16^ Initially, local tissue-resident inflammatory cells are activated, followed by blood brain barrier breakdown, recruitment of circulating pro-inflammatory immune cells, and ultimately activation of tissue repair and anti-inflammatory mechanisms. In aSAH, cerebrospinal fluid (CSF) proinflammatory molecules are associated with vasospasm, delayed cerebral ischemia (DCI), and worse outcomes.^17–21^ While recent human studies on ICH have reported an initial proinflammatory phase involving central nervous system (CNS)-resident and circulating immune cells followed by an anti-inflammatory reparative phase,^22^ data on inflammatory processes within the CNS and clinical outcome in patients following ICH and aSAH remains sparse.

Prior studies investigating the inflammatory profile of CSF found variable associations between immune molecules and outcome and focused on ICH or aSAH individually, limiting our understanding of differences in inflammatory responses between the diseases.^23,24^ In addition, few studies include both plasma and CSF cytokine levels to compare inflammatory responses across tissue compartments. In this study, we sought to more fully characterize CNS inflammation after hemorrhagic stroke in humans by analyzing a panel of inflammatory molecules in biological samples collected within the first 72 hours after symptom onset. To achieve this aim, we compared CSF and plasma levels of various immune molecules; compared CSF levels between ICH, aSAH, and control patients; and investigated relationships between these inflammatory molecules and markers of hemorrhage severity.

## METHODS

### Study design

In this case-control study, 242 patients with spontaneous ICH or aSAH admitted to the Yale Neurosciences Intensive Care Unit (ICU) between 2014 and 2023 with plasma and/or CSF samples collected for the Yale Acute Brain Injury Biorepository were included. CSF was collected from patients who had external ventricular drains (EVDs) placed as part of their clinical care. Eighteen patients undergoing ventriculoperitoneal shunt placement for normal pressure hydrocephalus (NPH) were enrolled as controls, and CSF was collected during shunt placement. This study was approved by Yale School of Medicine IRB, and consent was obtained from patients or their legally authorized representative. Data reporting adheres to the STROBE guidelines for reporting of observational studies.

Blood and CSF from eligible patients were collected between presentation and 72 hours after symptom onset, and the first sample collected from each patient was included. Patients with plasma and CSF samples collected at the same time (n = 16 ICH patients, n = 17 aSAH patients) were included in our paired samples cohort.

### Immunologic Assays

Within 30 minutes of collection, blood and CSF samples were centrifuged, the supernatant was divided into aliquots, and samples were frozen at –80°C. A panel of immune molecules including interleukin-6 (IL-6), granulocyte-colony stimulating factor (G-CSF), IL-8, vascular endothelial growth factor (VEGF), C-C motif chemokine ligand-2 (CCL2), IL-1β, IL-4, macrophage inflammatory protein-1 alpha (MIP-1α), IL-10, tumor necrosis factor alpha (TNF-α), and IL-17 were analyzed using Cytometric Bead Array (BD Biosciences, Franklin Lakes, NJ), a multiplex bead-based immunoassay. Based on prior work in our laboratory showing that VEGF was not detectable in the plasma of hemorrhagic stroke patients at these timepoints (unpublished data), VEGF was not measured in plasma samples. The lower limit of detection for each immune molecule was the concentration of the lowest standard for that assay.

### Independent Variables

Independent variables were immune molecules above the lower limit of detection, namely IL-6, CCL2, IL-8, G-CSF, and VEGF. ICU length of stay was extracted from the electronic health record. For patients with multiple ICU admissions during the same hospitalization, the sum of all ICU days was considered their ICU length of stay.

### Clinical and Neuroimaging Variables

Demographics, medical history, and hospitalization data was obtained from chart review and follow-up interviews with patients and their families. Clinical and radiographic scores including Glasgow Coma Scale (GCS) scores for all patients and Hunt and Hess scores and Modified Fisher Scale scores for aSAH patients, were extracted from patient charts by trained raters (JY, RC). If disagreement or uncertainty existed between raters, the scores were adjudicated by a board certified neurointensivist (JMB, JK). For ICH patients, imaging was reviewed by a board certified neurointensivist (JMB) who calculated ICH volumes using the ABC/2 method,^25^ determined whether IVH was present, and classified ICH location. ICH score^10^ was then calculated using the above data by a trained rater (JY). Patients with primary IVH not attributable to a vascular etiology were characterized as ICH. Patients with a ruptured aneurysm, even if the hemorrhage was largely intraparenchymal, were characterized as aSAH.

### Statistical Analysis

Non-normally distributed concentrations of immune molecules were log transformed. In our paired samples cohort, paired t-test was used to compare log CSF and plasma concentrations of the immune molecules. For CSF samples, ANOVA was used to compare log CSF immune molecule concentrations between control, ICH, and aSAH samples. Tukey post hoc test was used to determine differences between groups. Linear regression was used to evaluate for significant associations between log CSF immune molecule concentrations and hemorrhage severity features. There was no missing clinical or demographic data. For samples in which there was missing immune molecule data, those samples were excluded from analysis only of the missing molecule. There was no a priori sample size calculation as this was an analysis of previously collected samples. Based on the 72 CSF samples collected, the study had 80% power to detect a 13% difference or a 95% power to detect a 20% difference in log-transformed immune molecules between control (n = 18), ICH (n = 21), and aSAH (n = 51) samples using a one-way ANOVA with an alpha = 0.05. R software (version 4.3.1) was used for all data management and statistical analysis.

## RESULTS

Plasma and/or CSF samples from 260 ICH (n = 188), aSAH (n = 54), and control (n = 18) patients were analyzed. Patients had a median age of 67 years (interquartile range [IQR] 18-95), 51.5% were male, 92.7% were non-Hispanic, and 76.2% were Caucasian (Table 1). In ICH and aSAH patients, the median time from symptom onset to sample collection was 3 [IQR 2] days for CSF and 1 [IQR 2] days for plasma. The measured levels of each inflammatory molecule and lower limits of detection are shown in Supplemental Table 1 for plasma samples and Supplemental Table 2 for CSF samples.

**Table 1.** Baseline Demographics (All subjects)

|  | Control<br>(N=18) | ICH<br>(N=188) | SAH<br>(N=54) | All Subjects<br>(N=260) |
| --- | --- | --- | --- | --- |
| **Sample type<sup>†</sup>** |  |  |  |  |
| Plasma | NA | 181 | 21 | 202 |
| CSF | 18 | 21 | 51 | 90 |
| **Age** |  |  |  |  |
| Mean (SD) | 77.3 (4.21) | 67.1 (14.3) | 57.8 (14.2) | 65.9 (14.6) |
| Median [Min, Max] | 76.5 [71.0, 88.0] | 68.0 [19.0, 95.0] | 58.0 [18.0, 90.0] | 67.0 [18.0, 95.0] |
| **Sex** |  |  |  |  |
| Female | 5 (27.8%) | 84 (44.7%) | 37 (68.5%) | 126 (48.5%) |
| Male | 13 (72.2%) | 104 (55.3%) | 17 (31.5%) | 134 (51.5%) |
| **Ethnicity** |  |  |  |  |
| Hispanic | 0 (0%) | 7 (3.7%) | 10 (18.5%) | 17 (6.5%) |
| Non-Hispanic | 18 (100%) | 179 (95.2%) | 44 (81.5%) | 241 (92.7%) |
| Unknown | 0 (0%) | 2 (1.1%) | 0 (0%) | 2 (0.8%) |
| **Race** |  |  |  |  |
| White or Caucasian | 18 (100%) | 144 (76.6%) | 36 (66.7%) | 198 (76.2%) |
| Black or African American | 0 (0%) | 30 (16.0%) | 5 (9.3%) | 35 (13.5%) |
| American Indian | 0 (0%) | 1 (0.5%) | 0 (0%) | 1 (0.4%) |
| Asian | 0 (0%) | 3 (1.6%) | 3 (5.6%) | 6 (2.3%) |
| Other | 0 (0%) | 6 (3.2%) | 6 (11.1%) | 12 (4.6%) |
| Unknown | 0 (0%) | 4 (2.1%) | 4 (7.4%) | 8 (3.1%) |
| ICH, intracerebral hemorrhage; SAH, subarachnoid hemorrhage; SD, standard deviation; CSF, cerebrospinal fluid; <sup>†</sup>33 patients had both CSF and plasma samples collected |  |  |  |  |

### Inflammatory molecules are elevated in the CSF compared to plasma

CCL2, IL-6, G-CSF, IL-8, and VEGF were detectable in CSF of patients within 72 hours after symptom onset of ICH and aSAH. These proinflammatory molecules were present in higher concentrations in the CSF compared to the plasma (Figure 1, p<0.05 for all comparisons).

**Figure 1.**
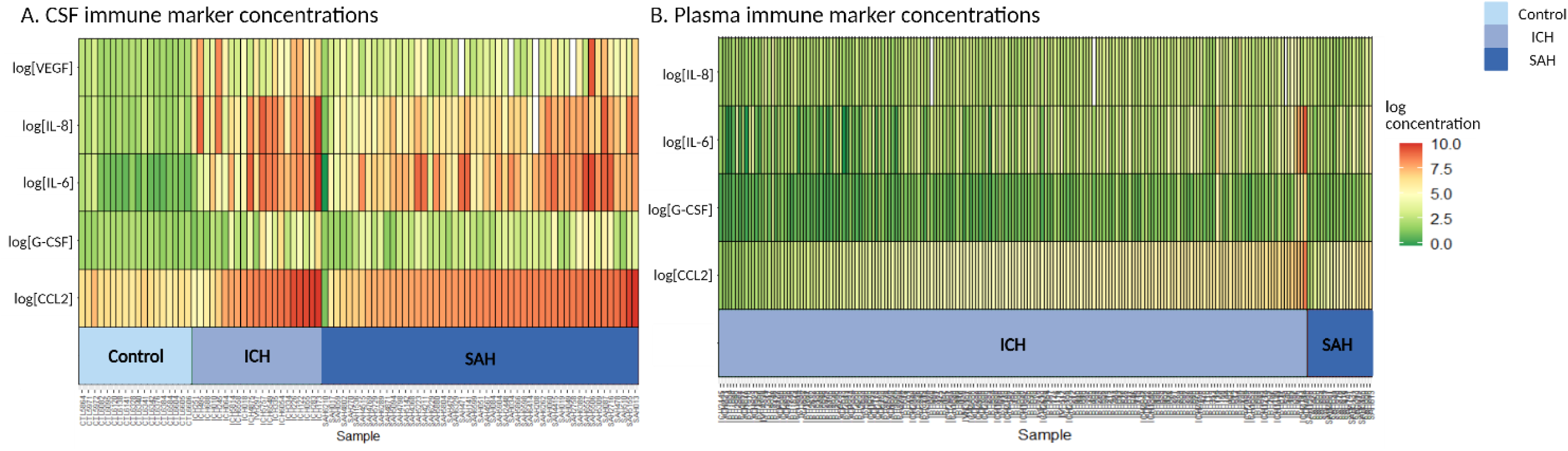
– Comparison of log immune molecule concentrations between CSF and plasma. Heat maps demonstrate higher pro-inflammatory immune molecule in CSF compared to plasma at early timepoints.

In 16 ICH and 17 aSAH patients, paired CSF and plasma samples collected at the same timepoint were available. Mean log concentrations of all detectable inflammatory markers were significantly higher in CSF of ICH patients compared to plasma samples, including CCL2 (CSF 8.16pg/mL vs plasma 4.73pg/mL, p<0.01), IL-6 (CSF 6.53pg/mL vs plasma 3.49pg/mL, p<0.01), G-CSF (CSF 2.95pg/mL vs plasma 1.89pg/mL, p<0.01), and IL-8 (CSF 7.19pg/mL vs plasma 3.96pg/mL, p<0.01; Figure 2A). Additionally, mean log concentrations of these cytokines were also significantly higher in SAH patient CSF compared to plasma samples, including CCL2 CSF 8.36pg/mL vs plasma 4.71pg/mL, p<0.01), IL-6 (CSF 7.07pg/mL vs plasma 3.10pg/mL, p<0.01), G-CSF (CSF 2.79pg/mL vs plasma 1.69pg/mL, p<0.01), and IL-8 (CSF 6.72pg/mL vs plasma 2.57pg/mL, p<0.01; Figure 2B).

**Figure 2.**
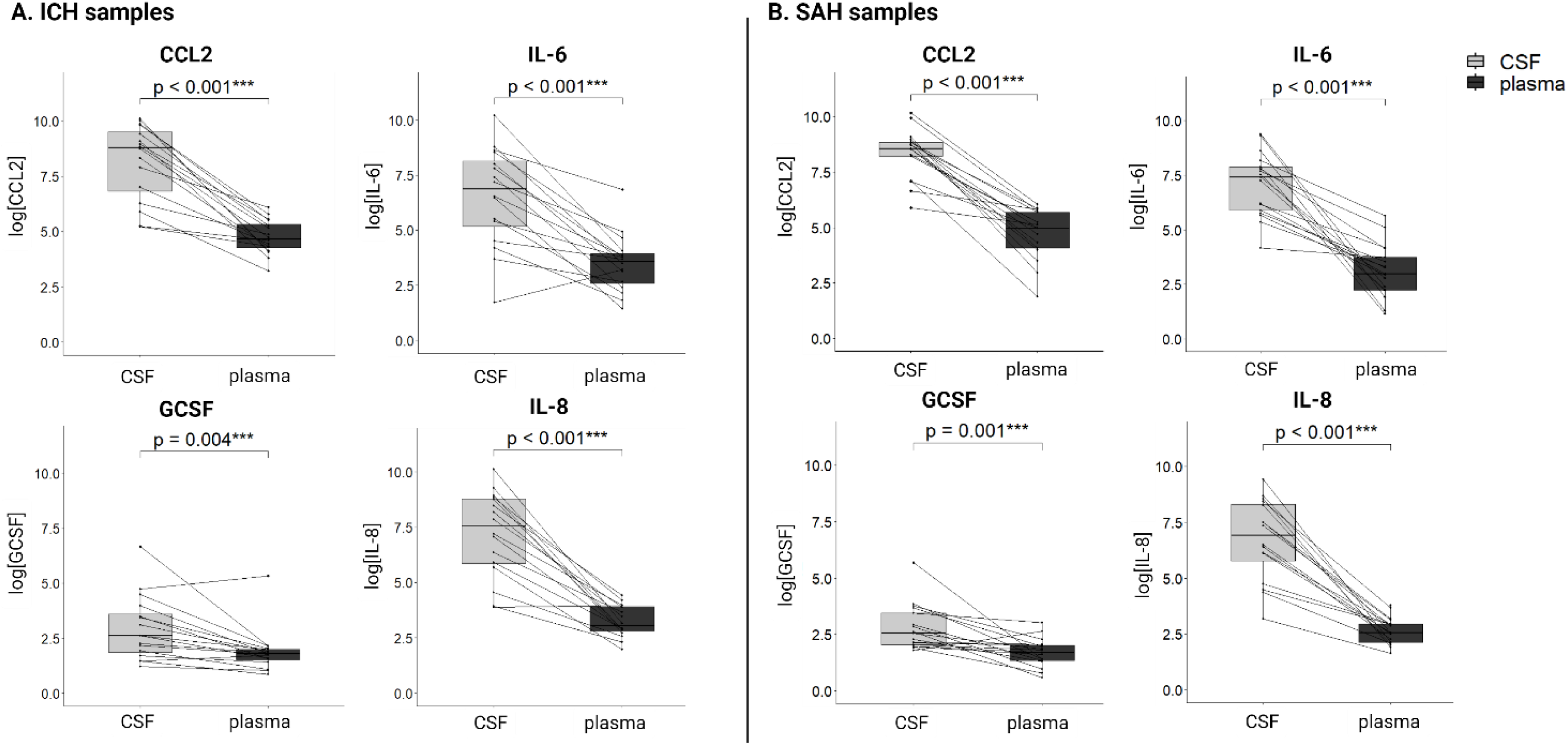
– Comparison of log immune molecule concentrations between CSF and plasma in paired patient samples. In paired samples collected from the same patient at the same timepoint, immune molecule concentrations are higher in the CSF compared to the plasma.

### Differences in CSF inflammatory molecules across hemorrhagic stroke type

Compared to CSF collected from controls, patients with both hemorrhagic stroke subtypes had significantly higher CSF concentrations of CCL2 (controls 6.50pg/mL vs ICH 8.24pg/mL vs mean aSAH 8.19pg/mL, p<0.01), IL-6 (controls 1.32pg/mL vs ICH 6.89pg/mL vs aSAH 7.29pg/mL, p<0.01), G-CSF (controls 2.04pg/mL vs ICH 3.46pg/mL vs SAH 3.12pg/mL, p<0.01), and IL-8 (controls 2.23pg/mL vs ICH 7.31pg/mL vs aSAH 6.49pg/mL, p<0.01) (all results reported in mean log concentrations, Figure 3A-D). Patients with hemorrhagic stroke had significantly higher concentrations of CSF VEGF compared to controls, and ICH patients had significantly higher CSF VEGF than aSAH patients (controls 2.25pg/mL vs ICH 5.38 vs SAH 4.02pg/mL, p<0.01; Figure 3E).

**Figure 3.**
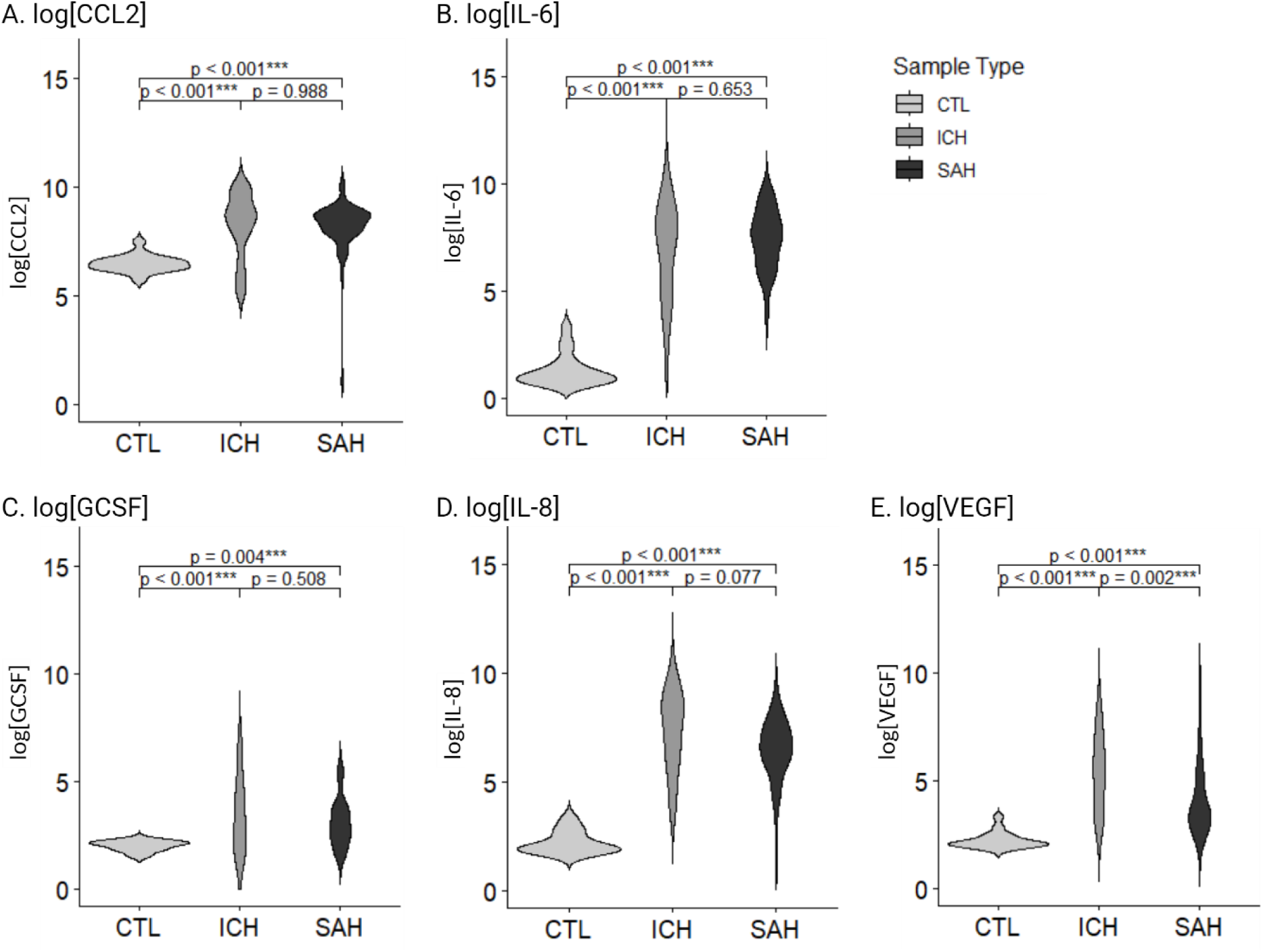
– Comparison of CSF immune molecule concentrations between control, ICH, and SAH patients. Within the CSF, immune molecule concentrations differ between control, ICH, and SAH patients.

### Association of inflammatory molecules and features of severity

Patients included in our study demonstrated features associated with severe neurological injury (Table 2).

**Table 2.** Patient Characteristics (CSF samples)

|  | ICH<br>(N=21) | SAH<br>(N=51) | All Samples<br>(N=72) |
| --- | --- | --- | --- |
| **Admission Glasgow Coma Scale score** |  |  |  |
| Mean (SD) | 5.20 (3.21) | 9.37 (4.68) | 8.20 (4.69) |
| Median [Min, Max] | 3.50 [3.00, 15.0] | 10.0 [3.00, 15.0] | 7.00 [3.00, 15.0] |
| **ICU Length of Stay (days)** |  |  |  |
| Mean (SD) | 13.1 (8.22) | 17.8 (9.28) | 16.5 (9.19) |
| Median [Min, Max] | 13.2 [0.708, 35.1] | 15.1 [4.25, 55.6] | 14.8 [0.708, 55.6] |
| **Hunt Hess Score** |  |  |  |
| Mean (SD) | NA | 3.24 (1.14) | 3.24 (1.14) |
| Median [Min, Max] | NA | 3.00 [2.00, 5.00] | 3.00 [2.00, 5.00] |
| **Modified Fisher Scale Score** |  |  |  |
| Mean (SD) | NA | 3.65 (0.594) | 3.65 (0.594) |
| Median [Min, Max] | NA | 4.00 [2.00, 4.00] | 4.00 [2.00, 4.00] |
| **ICH Score** |  |  |  |
| Mean (SD) | 2.89 (0.676) | NA | 2.89 (0.676) |
| Median [Min, Max] | 3.00 [2.00, 4.00] | NA | 3.00 [2.00, 4.00] |
| **ICH Volume (mL)** |  |  |  |
| Mean (SD) | 35.3 (30.1) | NA | 35.3 (30.1) |
| Median [Min, Max] | 31.7 [0, 102] | NA | 31.7 [0, 102] |
| ICH: intracerebral hemorrhage; SAH: subarachnoid hemorrhage; SD: standard deviation; ICU: intensive care unit |  |  |  |

In CSF collected from ICH patients, a lower log concentration of CCL2 was associated with a larger ICH volume (p=0.037). There was no association between ICH volume and any of the other inflammatory molecules. There were no significant associations between CSF concentrations of the measured inflammatory molecules and admission GCS, ICU length of stay, or ICH score (all p>0.05; Table 3).

**Table 3.**
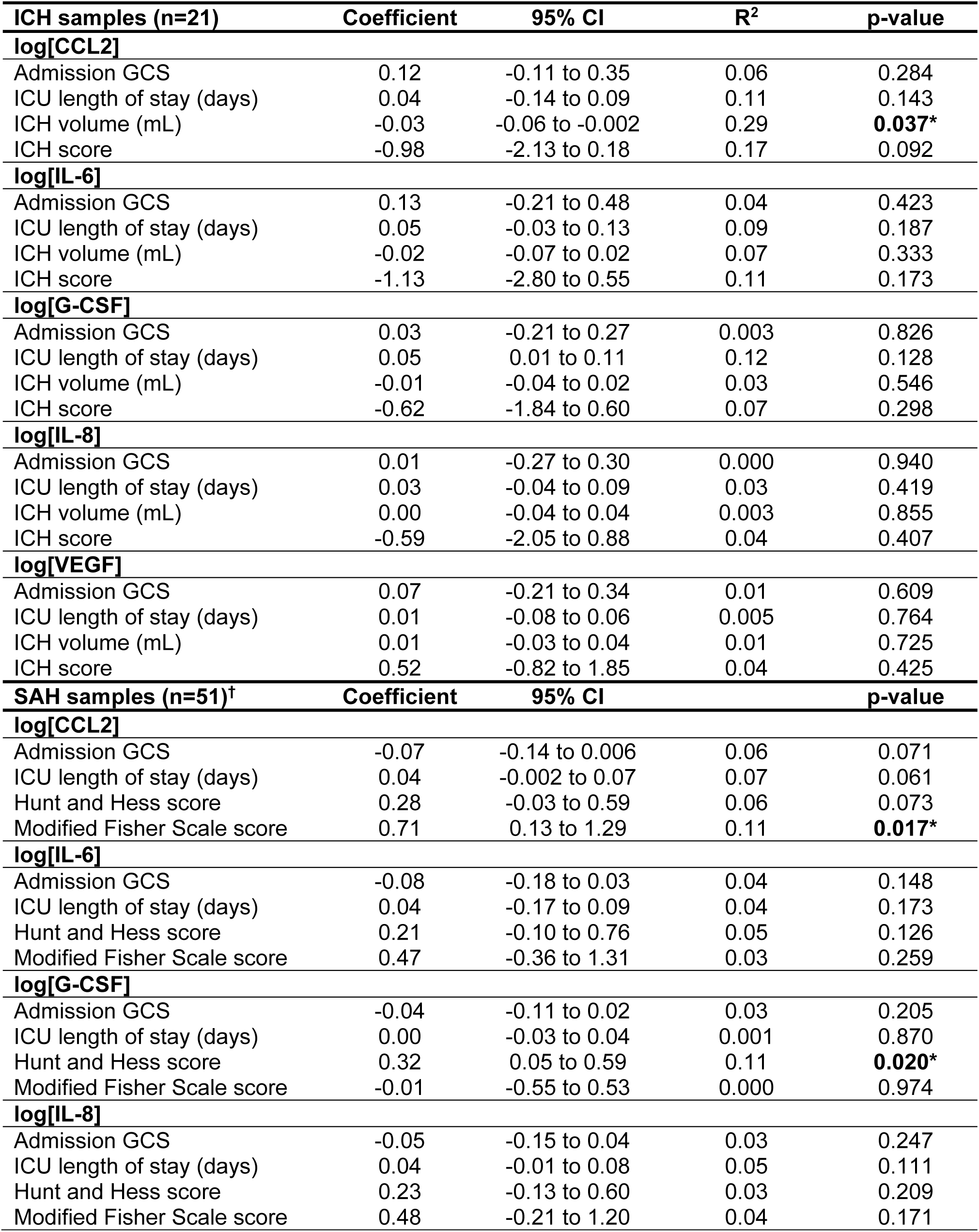
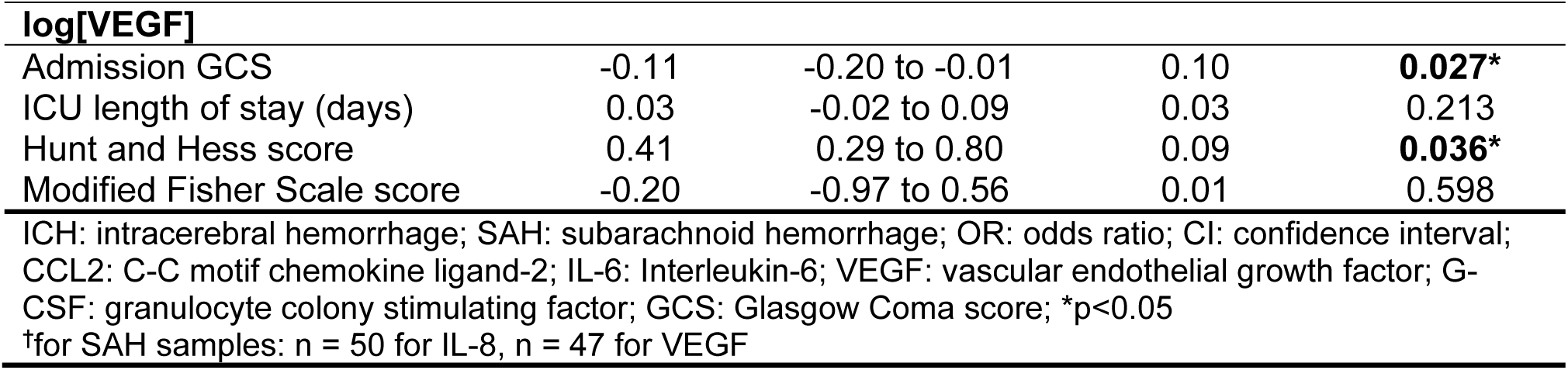
Association of CSF inflammatory molecule concentrations with severity factors (univariate analysis)

For aSAH patients, higher log concentration of VEGF was associated with a lower admission GCS score (p=0.027). Higher log concentrations of G-CSF and VEGF were associated with higher Hunt and Hess scores (G-CSF p=0.020, VEGF p=0.036). Additionally, higher log concentration of CCL2 was associated with higher modified Fisher Scale scores (p=0.017). There were no associations between CSF inflammatory molecules and ICU length of stay in aSAH patients (all p>0.05; Table 3).

## DISCUSSION

Inflammation occurs in the CNS after hemorrhagic stroke, and it is possible to detect inflammatory molecules in the CSF of these patients. In this study, we have shown that concentrations of several proinflammatory molecules are higher within the CSF as compared to the plasma of patients in the first 3 days after hemorrhage. These proinflammatory molecules were higher in the CSF in the hemorrhagic stroke patients overall, and within individual patients for whom we had both plasma and CSF samples collected on the same day. Additionally, within the CSF, proinflammatory molecules were higher in ICH and aSAH patients compared to controls and VEGF was significantly higher in ICH patients compared to aSAH patients.

Several clinical and radiographic features are associated with more severe presentation or worse outcomes after ICH and aSAH including admission GCS score and ICU length of stay for both groups,^10,26–28^ Hunt and Hess score and modified Fisher Scale score for aSAH patients,^6,29,30^ and ICH score and initial ICH volume for ICH patients.^10^ In this hypothesis-generating analysis, we found that several of these features of disease severity were associated with CSF inflammatory molecule concentrations. Most notably, higher CSF VEGF and G-CSF were associated with higher Hunt and Hess scores and lower admission GCS, higher VEGF was associated with lower admission GCS score, and higher CCL2 was associated with higher modified Fisher Scale score for aSAH patients. Interestingly, higher CSF CCL2 in this early time window was associated with a smaller ICH volume. We did not find any significant associations between IL-6 or IL-8 concentrations and severity features.

CCL2, a monocyte chemoattractant, IL-8, a neutrophil chemoattractant, and IL-6, a multifunctional proinflammatory cytokine that stimulates production of acute phase proteins, are involved in the early proinflammatory phase that occurs within the CSF after hemorrhage and likely play a role in the recruitment of circulating immune cells to the CNS. In addition to these chemoattractant molecules, cell adhesion molecules responsible for the recruitment of circulating immune cells into the CNS have also been shown to be elevated in the CSF of ICH patients.^24^ We hypothesize that the presence of these inflammatory molecules early after hemorrhage is not necessarily detrimental, but that increased levels or prolonged elevation beyond the acute period may worsen outcomes and prevent repair mechanisms.

Our findings of elevated proinflammatory molecules early after hemorrhagic stroke is consistent with the prior literature. CCL2 has previously been shown to be higher in the CSF compared to the plasma of aSAH patients within the first 1 to 2 days after hemorrhage, and has also been reported to be associated with elevated IL-6 and correlated with higher risk of vasospasm.^31–33^ Although we did not assess the relationship between CSF CCL2 and vasospasm in this study, we found an association between early CSF CCL2 and modified Fisher Scale score, a marker of patients at risk for vasospasm and DCI. In ICH patients, CSF CCL2 has been shown to be elevated within the first 1 to 2 days after hemorrhage, and was associated with ICH volume and volume of perihematomal edema.^34^ In contrast, we find an association of CSF CCL2 with smaller ICH volume, although notably in our study ICH volume did not include the volume of IVH.

Prior studies of CSF IL-6 in hemorrhagic stroke have focused on aSAH patients with studies showing elevated CSF IL-6 compared to patient plasma and control CSF, as well as elevated CSF IL-6 in patients with worse outcome, vasospasm, and DCI.^17,18,31,35–40^ In ICH patients, elevated early CSF IL-6 is associated with hematoma expansion, larger hematoma volume, worse outcomes, and following intraventricular administration of tissue plasminogen activator.^34,41^ Our findings are consistent with prior findings of elevated early CSF IL-6 compared to plasma levels and control CSF, but in our small cohort of patients we were not able to assess outcomes or complications, and we did not find an association with disease severity.

CSF IL-8 has been less well studied after hemorrhagic stroke. IL-8 has been found to be elevated in the CSF of aSAH patients in the first week after hemorrhage, along with IL-6.^31,42^ In ICH patients, CSF IL-8 levels 1 week post-bleed were found to be associated with poor admission GCS and IVH volume.^34^ In the current study, we found high levels of IL-8 early after hemorrhage in CSF of both ICH and aSAH patients compared to plasma levels and compared to control CSF, although we found no association with disease severity.

In the current study, we report increased levels of VEGF and G-CSF within the CSF after hemorrhagic stroke, and we find associations between these molecules and severity features. There is limited prior investigation of these molecules after hemorrhagic stroke. In aSAH patients, CSF VEGF was elevated compared to patient plasma and control CSF, but was not associated with outcome or vasospasm.^31^ In ICH, VEGF has been suggested to play an anti-inflammatory role, as it is induced by prostaglandin E2 in human macrophages stimulated with ICH damage-associated molecular patterns.^22^ Interestingly, CSF VEGF is the only molecule in our study which differed between ICH and aSAH patients, and we showed that it was higher in the CSF of ICH patients despite its classic role in angiogenesis. VEGF may play a dual role in the CNS, as it has been shown to be involved in blood brain barrier permeability, but also appears to play an anti-inflammatory role and is involved in vessel repair.^43,44^

G-CSF within the CSF of hemorrhagic stroke patients has not been previously investigated, but in a rat model of ICH it was found to be neuroprotective and associated with increased VEGF expression after hemorrhage.^45^ G-CSF, which is primarily produced by monocytes, is classically involved in the proliferation and differentiation of granulocytes, but has also been shown to be neuroprotective and play a role in plasticity in ischemic stroke.^46–48^ It is intriguing that early CSF levels of VEGF and G-CSF are associated with features of severity in our study, as these molecules have been previously shown to have some neuroprotective and anti-inflammatory effects, as opposed to CCL2, IL-6, and IL-8 which are predominantly proinflammatory, but this is perhaps not surprising for VEGF which appears to have a time-dependent role in the CNS after injury.

Our study has several limitations and is meant to be hypothesis generating. First, we only collected samples within the first 3 days after ictus, limiting our investigation to the acute inflammatory response. We believe that these early proinflammatory molecules are most likely to be associated with initial disease severity features. Future work with longitudinal CSF samples from hemorrhagic stroke patients will be needed to further understand the temporal changes in these and other inflammatory molecules throughout the disease course. Secondly, we are only able to collect CSF samples from patients with EVDs placed as part of their clinical care, and as such we only have data on more severely affected ICH and aSAH patients. While this is only a subset of ICH and aSAH patients, these patients are at highest risk for poor outcomes so a better understanding their disease course and developing therapeutics for this population is likely to have a substantial impact on their outcome. Our small sample size limits our statistical power but has generated several hypotheses for ongoing investigation into the role of CNS inflammation after hemorrhagic stroke. Finally, our analysis was limited to 11 immune molecules, and thus was not a comprehensive study of all factors that may be associated with hemorrhagic brain injury and outcomes.

Measurement of inflammatory molecules within the CNS is critically important to characterize the inflammatory cascade that occurs after hemorrhagic stroke, and evaluation of inflammatory molecules in the blood is insufficient. By focusing on the temporal trajectory of CNS inflammatory molecules, we aim to better define the kinetics of inflammation in the CNS following hemorrhage. While there is overlap between the inflammation that occurs following ICH and aSAH, given their distinct etiologies there are also likely to be important and informative differences. In this study we have begun to explore the relationship between early inflammatory molecules and disease severity. Future studies on the relationship with clinical outcomes will be important to further define potential therapeutic strategies.

## Data Availability

A deidentified version of the data is available upon request.

## Non-standard Abbreviations and Acronyms

ICH: intracerebral hemorrhage
aSAH: aneurysmal subarachnoid hemorrhage
DCI: delayed cerebral ischemia
CSF: cerebrospinal fluid
EVD: external ventricular drain
CCL2: C-C motif chemokine ligand-2
IL-6: interleukin-6
G-CSF: granulocyte-colony stimulating factor
IL-8: interleukin-8
VEGF: vascular endothelial growth factor
CNS: central nervous system
NPH: normal pressure hydrocephalus
IL-1β: interleukin-1 beta
IL-4: interleukin-4
MIP-1α: macrophage inflammatory protein-1 alpha
IL-10: interleukin-10
TNF-α: tumor necrosis factor alpha
IL-17: interleukin-17
ICU: Intensive Care Unit
GCS: Glasgow Coma Scale
IQR: interquartile range
ANOVA: analysis of variance

## ACKNOWLEDGEMENTS

The authors would like to thank Manali Phadke for assistance with statistical analysis. Figures were created with BioRender.com

## REFERENCES

1. Pinho J, S CA, Araujo JM, Amorim JM, Ferreira C. Intracerebral hemorrhage outcome: a comprehensive update. Journal of the Neurologic Sciences. 2019;398:54–66.

2. Nieuwkamp DJ, Setz LE, Algra A, Linn FHH, deRooij NK, Rinkel GJE. Changes in case fatality of aneurysmal subarachnoid haemorrhage over time, according to age, sex, and region: a meta-analysis. Lancet Neurology. 2009;8:365–342.

3. Rinkel GJE, Algra A. Long-term outcomes of patients with aneurysmal subarachnoid haemorrhage. Lancet Neurology. 2011;10:349–356.

4. Lovelock CE, Rinkel GJE, Rothwell PM. Time trends in outcome of subarachnoid hemorrhage: population-based study and systematic review. Neurology. 2010;74:1494–1501.

5. Sadaf H, Desai VR, Misra V, Golanov E, Hegde ML, Villapol S, Karmonik C, Regnier-Golanov A, Sayenko D, Horner PJ, et al. A contemporary review of therapeutic and regenerative management of intracerebral hemorrhage. Ann Clin Transl Neurol. 2021;8:2211–2221. doi: 10.1002/acn3.51443

6. Claassen J, Bernardini GL, Kreiter K, Bates J, Du YE, Copeland D, Connolly ES, Mayer SA. Effect of cisternal and ventricular blood on risk of delayed cerebral ischemia after subarachnoid hemorrhage: the Fisher scale revisited. Stroke. 2001;32:2012–2020.

7. Oppong MD, Gembruch O, Herten A, Frantzev R, Chihi M, Dammann P, El Hindy N, Forsting M, Sure U, Jabbarli R. Intraventricular hemorrhage caused by subarachnoid hemorrhage: does the severity matter? World Neurosurgery. 2018;111:e693–e702.

8. Chan E, Anderson CS, Wang X, Arima H, Saxena A, Moullaali TJ, Heeley E, Delcourt C, Wu G, Wang J, et al. Significance of intraventricular hemorrhage in acute intracerebral hemorrhage: intensive blood pressure reduction in acute cerebral hemorrhage trial results. Stroke. 2015;46:653–658.

9. Hanley DF. Intraventricular hemorrhage: severity factor and treatment target in spontaneous intracerebral hemorrhage. Stroke. 2009;40:1533–1538. doi: 10.1161/STROKEAHA.108.535419

10. Hemphill III JC, Bonovich DC, Besmertis L, Manley GT, Johnston SC. The ICH Score: A simple, reliable grading scale for intracerebral hemorrhage. Stroke. 2001;32:891–897.

11. Agnihotri S, Czap A, Staff I, Fortunato G, McCullough L. Peripheral leukocyte counts and outcomes after intracerebral hemorrhage. Journal of Neuroinflammation. 2011;8:160.

12. Kwan K, Arapi O, Wagner KE, Schneider J, Sy HL, Ward MF, Sison CP, Li C, Eisenberg MB, Chalif D, et al. Cerebrospinal fluid macrophage migration inhibitory factor: a potential predictor of cerebral vasospasm and clinical outcome after aneurysmal subarachnoid hemorrhage. J Neurosurg. 2019:1–6. doi: 10.3171/2019.6.JNS19613

13. Liu S, Liu X, Chen S, Xiao Y, Zhuang W. Neutrophil-lymphocyte ratio predicts the outcome of intracerebral hemorrhage: A meta-analysis. Medicine. 2019;98:e16211.

14. Hallevi H, Walker KC, Kasam M, Bornstein N, Grotta JC, Savitz SI. Inflammatory response to intraventricular hemorrhage: time course, magnitude and effect of t-PA. J Neurol Sci. 2012;315:93–95. doi: 10.1016/j.jns.2011.11.019

15. Walsh KB, Sekar P, Langefeld CD, Moomaw CJ, Elkind MS, Boehme AK, James ML, Osborne J, Sheth KN, Woo D, et al. Monocyte Count and 30-Day Case Fatality in Intracerebral Hemorrhage. Stroke. 2015;46:2302–2304. doi: 10.1161/STROKEAHA.115.009880

16. Askenase MH, Sansing LH. Stages of the Inflammatory Response in Pathology and Tissue Repair after Intracerebral Hemorrhage. Semin Neurol. 2016;36:288–297. doi: 10.1055/s-0036-1582132

17. Wu W, Guan Y, Zhao G, Fu XJ, Guo TZ, Liu YT, Ren XL, Wang W, Liu HR, Li YQ. Elevated IL-6 and TNF-alpha Levels in Cerebrospinal Fluid of Subarachnoid Hemorrhage Patients. Mol Neurobiol. 2016;53:3277–3285. doi: 10.1007/s12035-015-9268-1

18. Lenski M, Huge V, Briegel J, Tonn JC, Schichor C, Thon N. Interleukin 6 in the Cerebrospinal Fluid as a Biomarker for Onset of Vasospasm and Ventriculitis After Severe Subarachnoid Hemorrhage. World Neurosurg. 2017;99:132–139. doi: 10.1016/j.wneu.2016.11.131

19. Takizawa T, Tada T, Kitazawa K, Tanaka Y, Hongo K, Kameko M, Uemura KI. Inflammatory cytokine cascade released by leukocytes in cerebrospinal fluid after subarachnoid hemorrhage. Neurol Res. 2001;23:724–730. doi: 10.1179/016164101101199243

20. Mohme M, Sauvigny T, Mader MM, Schweingruber N, Maire CL, Runger A, Ricklefs F, Regelsberger J, Schmidt NO, Westphal M, et al. Immune Characterization in Aneurysmal Subarachnoid Hemorrhage Reveals Distinct Monocytic Activation and Chemokine Patterns. Transl Stroke Res. 2020;11:1348–1361. doi: 10.1007/s12975-019-00764-1

21. Yu F, Saand A, Xing C, Lee JW, Hsu L, Palmer OP, Jackson V, Tang L, Ning M, Du R, et al. CSF lipocalin-2 increases early in subarachnoid hemorrhage are associated with neuroinflammation and unfavorable outcome. J Cereb Blood Flow Metab. 2021;41:2524–2533. doi: 10.1177/0271678X211012110

22. Askenase MH, Goods BA, Beatty HE, Steinschneider AF, Velazquez SE, Osherov A, Landreneau MJ, Carroll SL, Tran TB, Avram VS, et al. Longitudinal transcriptomics define the stages of myeloid activation in the living human brain after intracerebral hemorrhage. Science Immunology. 2021;6:eabd6279.

23. Chaudhry SR, Stoffel-Wagner B, Kinfe TM, Guresir E, Vatter H, Dietrich D, Lamprecht A, Muhammad S. Elevated Systemic IL-6 Levels in Patients with Aneurysmal Subarachnoid Hemorrhage Is an Unspecific Marker for Post-SAH Complications. Int J Mol Sci. 2017;18. doi: 10.3390/ijms18122580

24. Kraus J, Gerriets T, Leis S, Stolz E, Oschmann P, Heckmann JG. Time course of VCAM-1 and ICAM-1 in CSF in patients with basal ganglia haemorrhage. Acta Neurol Scand. 2007;116:49–55. doi: 10.1111/j.1600-0404.2006.00790.x

25. Kothari RU, Brott T, Broderick JP, Barsan WB, Sauerbeck LR, Zuccarello M, Khoury J. The ABCs of measuring intracerebral hemorrhage volumes. Stroke. 1996;27:1304–1305.

26. Oshiro E, Walter K, Piantadosi S, Witham T, Tamargo R. A new subarachnoid hemorrhage grading system based on glasgow coma scale: a comparison with the hunt and hess and world federation of neurological surgeons scales in a clinical series. Neurosurgery. 1997;41:104–148.

27. Hammer A, Ranaie G, Erbguth F, Hohenhaus M, Wenzl M, Killer-Oberpfalzer M, Steiner HH, Janssen H. Impact of Complications and Comorbidities on the Intensive Care Length of Stay after Aneurysmal Subarachnoid Haemorrhage. Sci Rep. 2020;10:6228. doi: 10.1038/s41598-020-63298-9

28. Specogna AV, Turin TC, Patten SB, Hill MD. Hospital treatment costs and length of stay associated with hypertension and multimorbidity after hemorrhagic stroke. BMC Neurol. 2017;17:158. doi: 10.1186/s12883-017-0930-2

29. Frontera JA, Claassen J, Schmidt JM, Wartenberg KE, Temes R, Connolly ES, Macdonald RL, Mayer SA. Prediction of symptomatic vasospasm after subarachnoid hemorrhage: the modified Fisher scale. Neurosurgery. 2006;58:21–27.

30. Hunt WE, Hess RM. Surgical risk as related to time of intervention in the repair of intracranial aneurysms. Journal of Neurosurgery. 1968;28:14–20.

31. Al-Tamimi YZ, Bhargava D, Orsi NM, Teraifi A, Cummings M, Ekbote UV, Quinn AC, Homer-Vanniasinkam S, Ross S. Compartmentalisation of the inflammatory response following aneurysmal subarachnoid haemorrhage. Cytokine. 2019;123:154778. doi: 10.1016/j.cyto.2019.154778

32. Niwa A, Osuka K, Nakura T, Matsuo N, Watabe T, Takayasu M. Interleukin-6, MCP-1, IP-10, and MIG are sequentially expressed in cerebrospinal fluid after subarachnoid hemorrhage. J Neuroinflammation. 2016;13:217. doi: 10.1186/s12974-016-0675-7

33. Kim GH, Kellner CP, Hahn DK, Desantis BM, Musabbir M, Starke RM, Rynkowski M, Komotar RJ, Otten ML, Sciacca R, et al. Monocyte chemoattractant protein-1 predicts outcome and vasospasm following aneurysmal subarachnoid hemorrhage. J Neurosurg. 2008;109:38–43. doi: 10.3171/JNS/2008/109/7/0038

34. Ziai WC, Parry-Jones AR, Thompson CB, Sansing LH, Mullen MT, Murthy SB, Mould A, Nekoovaght-Tak S, Hanley DF. Early Inflammatory Cytokine Expression in Cerebrospinal Fluid of Patients with Spontaneous Intraventricular Hemorrhage. Biomolecules. 2021;11. doi: 10.3390/biom11081123

35. Fassbender K, Hodapp B, Rossol S, Bertsch T, Schmeck J, Schutt S, Fritzinger M, Horn P, Vajkoczy P, Kreisel S, et al. Inflammatory cytokines in subarachnoid haemorrhage: association with abnormal blood flow velocities in basal cerebral arteries. Journal of Neurology, Neurosurgery, and Psychiatry. 2001;70:534–537.

36. Mathiesen T, Andersson B, Loftenius A, von Holst H. Increased interleukin-6 levels in cerebrospinal fluid following subarachnoid hemorrhage. Journal of Neurosurgery. 1993;78:562–567.

37. Duris K, Neuman E, Vybihal V, Juran V, Gottwaldova J, Kyr M, Vasku A, Smrcka M. Early Dynamics of Interleukin-6 in Cerebrospinal Fluid after Aneurysmal Subarachnoid Hemorrhage. J Neurol Surg A Cent Eur Neurosurg. 2018;79:145–151. doi: 10.1055/s-0037-1604084

38. Schoch B, Regel JP, Wichert M, Gasser T, Volbracht L, Stolke D. Analysis of intrathecal interleukin-6 as a potential predictive factor for vasospasm in subarachnoid hemorrhage. Neurosurgery. 2007;60:828–836; discussion 828-836. doi: 10.1227/01.NEU.0000255440.21495.80

39. Ridwan S, Grote A, Simon M. Interleukin 6 in cerebrospinal fluid is a biomarker for delayed cerebral ischemia (DCI) related infarctions after aneurysmal subarachnoid hemorrhage. Sci Rep. 2021;11:12. doi: 10.1038/s41598-020-79586-3

40. Sarrafzadeh A, Schlenk F, Gericke C, Vajkoczy P. Relevance of cerebral interleukin-6 after aneurysmal subarachnoid hemorrhage. Neurocrit Care. 2010;13:339–346. doi: 10.1007/s12028-010-9432-4

41. Kramer AH, Jenne CN, Zygun DA, Roberts DJ, Hill MD, Holodinsky JK, Todd S, Kubes P, Wong JH. Intraventricular fibrinolysis with tissue plasminogen activator is associated with transient cerebrospinal fluid inflammation: a randomized controlled trial. J Cereb Blood Flow Metab. 2015;35:1241–1248. doi: 10.1038/jcbfm.2015.47

42. Osuka K, Watanabe Y, Suzuki C, Iwami K, Miyachi S. Sequential expression of neutrophil chemoattractants in cerebrospinal fluid after subarachnoid hemorrhage. J Neuroimmunol. 2021;357:577610. doi: 10.1016/j.jneuroim.2021.577610

43. Lange C, Storkebaum E, de Almodovar CR, Dewerchin M, Carmeliet P. Vascular endothelial growth factor: a neurovascular target in neurological diseases. Nat Rev Neurol. 2016;12:439–454. doi: 10.1038/nrneurol.2016.88

44. Zhang ZG, Zhang L, Jiang Q, Zhang R, Davies K, Powers C, Bruggen N, Chopp M. VEGF enhances angiogenesis and promotes blood-brain barrier leakage in the ischemic brain. J Clin Invest. 2000;106:829–838. doi: 10.1172/JCI9369

45. Liang S-D, Ma L-Q, Gao Z-Y, Zhuang Y-Y, Zhao Y-Z. Granulocyte colony-stimulating factor improves neurological function and nagiogenesis in intracerebral hemorrhage rats. European Review for Medical and Pharmacological Sciences. 2018;22:2005–2014.

46. Schneider A, Kuhn HG, Schabitz WR. A role for G-CSF (granulocyte-colony stimulating factor) in the central nervous system. Cell Cycle. 2005;4:1753–1757. doi: 10.4161/cc.4.12.2213

47. Modi J, Menzie-Suderam J, Xu H, Trujillo P, Medley K, Marshall ML, Tao R, Prentice H, Wu JY. Mode of action of granulocyte-colony stimulating factor (G-CSF) as a novel therapy for stroke in a mouse model. J Biomed Sci. 2020;27:19. doi: 10.1186/s12929-019-0597-7

48. Wallner S, Peters S, Pitzer C, Resch H, Bogdahn U, Schneider A. The Granulocyte-colony stimulating factor has a dual role in neuronal and vascular plasticity. Front Cell Dev Biol. 2015;3:48. doi: 10.3389/fcell.2015.00048

49. Mathiesen T, Edner G, Ulfarsson E, Andersson B. Cerebrospinal fluid interleukin-1 receptor antagonist and tumor necrosis factor-a following subarachnoid hemorrhage. Journal of Neurosurgery. 1997;87:215–220.

